# Accessibility, inclusivity, and implementation of COVID-19 clinical management guidelines early in the pandemic: a global survey

**DOI:** 10.1101/2021.03.31.21254680

**Authors:** Caitlin Pilbeam, Deborah Malden, Katherine Newell, Andrew Dagens, Kalynn Kennon, Melina Michelen, Nina Gobat, Louise Sigfrid

## Abstract

**Background:** With a rapidly changing evidence base, high-quality clinical management guidelines (CMGs) are key tools for aiding clinical decision making and increasing access to best available evidence-based care. A rapid review of COVID-19 CMGs found that most lacked methodological rigour, overlooked many at-risk populations, and had variations in treatment recommendations. Furthermore, social science literature highlights the complexity of implementing guidelines in local contexts where they were not developed and the resulting potential to compound health inequities. The aim of this study was to evaluate access to, inclusivity of, and implementation of Covid-19 CMGs in different settings.

**Methods:** A cross-sectional survey of clinicians worldwide from 15 June to 20 July 2020, to explore access to and implementation of Covid-19 CMGs and treatment and supportive care recommendations provided. Data on accessibility, inclusivity, and implementation of CMGs. were analyzed by geographic location.

**Results:** Seventy-six clinicians, from 27 countries responded, 82% from high-income countries, 17% from low-middle income countries. Most respondents reported access to Covid-19 CMG and confidence in implementation of these. However, many respondents, particularly from LMICs reported barriers to implementation, including limited access to treatments and equipment. Only 20% of respondents reported having access to CMGs covering care for children, 25% for pregnant women and 50% for older adults (>65 years). Themes emerging were for CMGs to include recommendations for different at-risk populations, and settings, include supportive care guidance, be readily updated as evidence emerges, and CMG implementation supported by training, and access to treatments recommended.

**Conclusion:** Our findings highlight important gaps in Covid-19 CMG development and implementation challenges during a pandemic, particularly affecting different at-risk populations and lower resourced settings., to improve access in evidence-based care recommendations during an emergency. The findings identifies an urgent need for an improved framework for CMG development, that is inclusive and adaptable to emerging evidence and considers contextual implementation support, to improve access to evidence-based care globally.

## Introduction

The Covid-19 pandemic is a global health emergency, with over 120 million cases and more than 2.7 million deaths worldwide (as of 30 March 2021) (1). Amidst considerable uncertainty, particularly at the beginning of the pandemic, clinicians looked toward national and international organisations, such as the World Health Organization (WHO), for clinical management guidance. For emerging infections, the main treatment early on is supportive care, such as oxygen, fluids, electrolyte balance and management of complications. To this end, expert bodies have produced evidence-based recommendations and clinical management guidelines (CMGs), for use by frontline clinicians. Developing evidence-based CMGs is resource intensive, and past studies show that in practice, many clinicians across the world may use CMGs produced by international organisations rather than local CMGs. However, recommendations produced in one context may not be directly applicable to other settings, with different risk factors, and available resources. (2,3)

CMGs are defined as “systematically developed statements to assist practitioner and patient decisions about appropriate healthcare for specific clinical circumstances.” (4) In practice, the implementation of CMGs aims to standardise best available evidence-based care and improve quality, effectiveness, and outcomes (5,6). In specific cases, evidence suggests that guidelines have measurable impacts on improving patient outcomes, morbidity, and mortality (7,8). However, literature also highlights the complexity of guideline implementation, some studies indicating disappointingly low adherence to guideline recommendations, and others highlighting how guidelines may also add to inequities experienced by disadvantaged groups (9–11). To be effective and context-appropriate, CMGs need to be evidence-based, accessible, high-quality, inclusive of the whole population, and their recommendations applicable to local context and resources. Recent studies highlight that CMGs produced during public health emergencies often fall short of the gold standard of guideline development (12,13).

A rapid review of Covid-19 guidelines in the early stages of the pandemic found that these CMGs were limited in their methodological rigour and lacked coverage of at-risk populations, including those with lower immune response due to age, illness, or medication (2). There were also wide variations in recommendations for empirical antimicrobial, antiviral, and experimental treatment. The majority of Covid-19 CMGs identified were produced in upper-middle and higher income countries, there were no Covid-19 CMG identified from lower income countries (LMICs) (2).

The knowledge of implementation of high consequence infectious diseases during public health emergencies in different resource settings is limited. The aim of this study is to address these gaps. his study is to our knowledge the first study presenting data on clinician’s experience of accessibility, inclusivity, and challenges in implementing Covid-19 CMGs including recommendations on treatment and supportive care early in the pandemic.

## Methodology

### Study design

We conducted a cross-sectional survey of frontline clinicians in primary and secondary care settings globally. The survey was open from 15 June to 20 July 2020.

The survey explored clinicians’ perception and experiences, including access to and challenges in implementation, of Covid-19 CMGs. Survey questions focused on clinicians’ confidence and recent experiences, including availability, quality, inclusivity, and implementation of Covid-19 CMGs.

### Data collection

The survey was developed by a multidisciplinary team of clinicians, epidemiologists, clinical and social science researchers with experience in outbreak research and response. Using convenience sampling, we disseminated the survey to a range of clinicians in different countries via clinical networks, including the Platform for European Preparedness Against (Re-) emerging Epidemics (PREPARE) (14), the International Severe Acute Respiratory and Emerging Infection Consortium (ISARIC) (15), and through informal clinical networks, in order to capture diverse experiences, priorities, and different context that may influence the use and perception of different guidelines and to gain a geographically representative sample. The survey was programmed onto the REDCap database (16,17), hosted at the University of Oxford, and circulated as an online link with two follow-up reminders to non-responders. The survey was disseminated via direct e-mails, newsletters, and social media for wide reach and inclusivity. The survey was designed to be brief to facilitate uptake by frontline clinicians, during the pandemic (Supplemental File 1).

### Data analysis

Most answers were structured as a seven point Likert scale response. Positive Likert scale responses (either agree or strongly agree) were coded as 1, and non-responses or negative responses (neither agree nor disagree, disagree, or strongly disagree) were coded as 0. Dichotomous responses of yes or no were coded as 1 or 0, respectively. Proportions were calculated whereby a total acceptability score was estimated as a percentage of interviewees providing a positive response (18). All analysis was conducted in R version 3.4.2. The data is presented in a narrative form. Income classifications are based on World Bank income brackets (19).

### Public and Patient Involvement

This study is part of a wider project to assess availability, quality, and inclusivity of CMGs for high consequence infectious diseases, including systematic reviews of Covid-19 CMGs. The study protocols and interpretations of the findings have been informed by patient groups.

### Ethics

Responses were anonymized, all participants consented to sharing of their anonymized data. This research was performed in accordance with the Declaration of Helsinki, and was given an exemption from ethical review by the Oxford Tropical Research Ethics Committee (OXTREC) on 16 April 2020.

## Results

Of the 76 respondents (51% male), caring for patients in 27 different countries, across six continents (Fig.1), most were doctors and 87% worked in a hospital, of which 18% worked in an intensive care unit (Table 1). Fifty-three percent of respondents reported having taken on more responsibility during the pandemic and 40% that they had to work in a different clinical role to support the Covid-19 response, with most supporting Covid-19 care, with some supporting research, public health or diagnostics, and 20% reported stepping up into a more senior clinical role.

**Figure 1.**
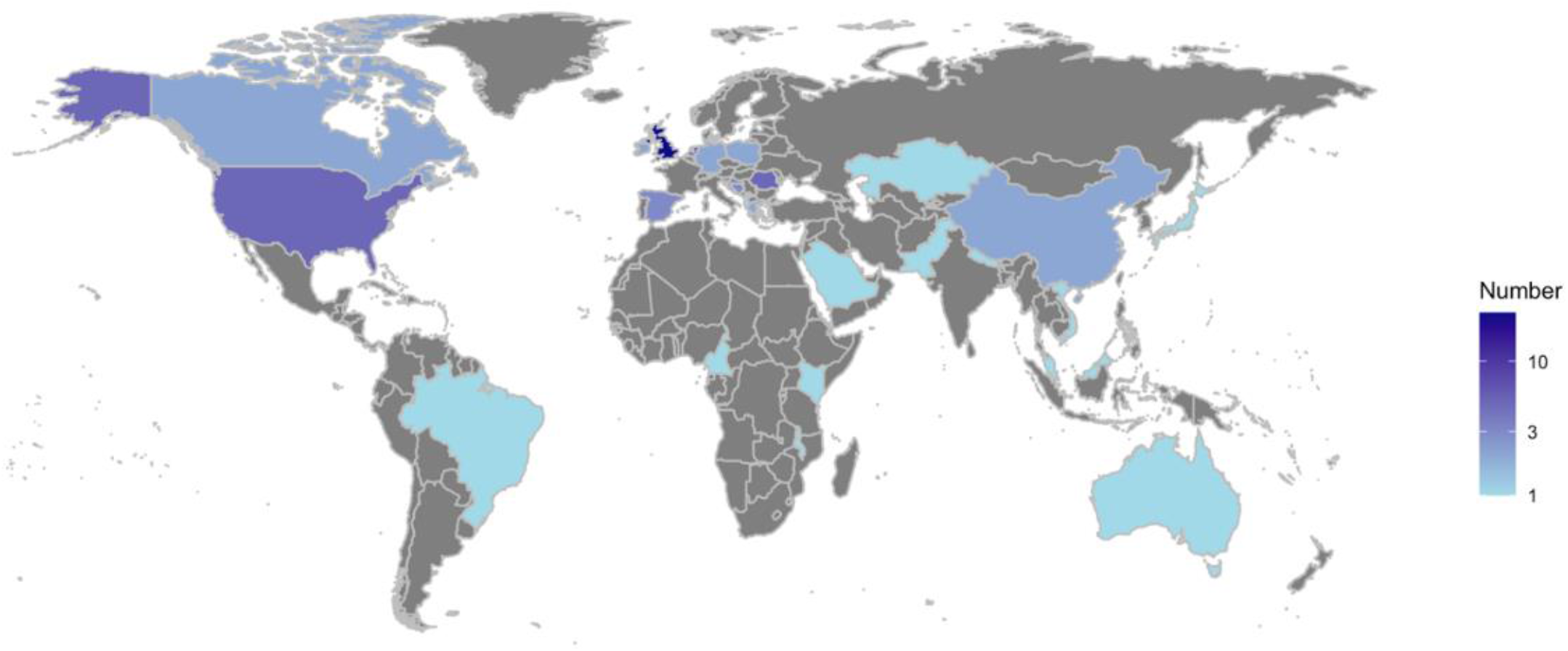
Geographic distribution of survey respondents. The map shows the number of respondents by country

**Figure 2.**
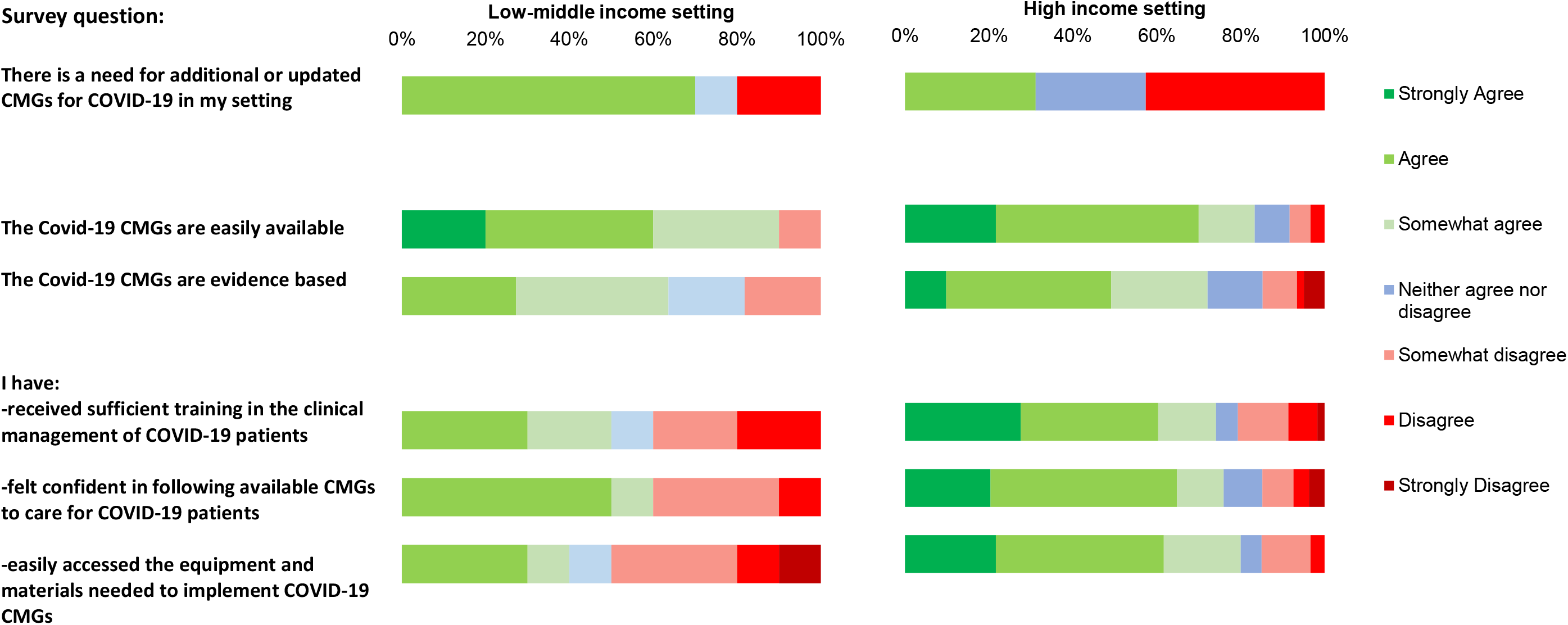
Accessibility, quality, and implementation of COVID-19 clinical management guidelines by setting. **Abbreviations:** CMGs: Clinical management guidelines

**Table 1:** Characteristics of survey respondents.

| Characteristics | Responses<br>n (%) |
| --- | --- |
| **Gender** |  |
| Male | 39 (51.3) |
| Female | 35 (46.1) |
| Not stated | 1 (1.3) |
| *Missing* | 1 (1.3) |
| **Age (years)** |  |
| 25-34 | 8 (10.5) |
| 35-44 | 23 (30.3) |
| 45-54 | 27 (35.5) |
| 55-64 | 15 (19.7) |
| 65-70 | 3 (3.9) |
| **Profession** |  |
| Senior medical doctor | 50 (65.8) |
| Junior medical doctor | 6 (7.9) |
| Nurse | 6 (7.9) |
| Other (undefined) | 11 (14.6) |
| **Region** |  |
| East Asia and Pacific | 6 (7.9) |
| Europe and Central Asia | 55 (72.4) |
| Latin America and Caribbean | 1 (1.3) |
| Middle East and North Africa | 1 (1.3) |
| South Asia | 2 (2.6) |
| Sub-Saharan Africa | 3 (3.9) |
| North America | 7 (9.2) |
| *Missing* | 1 (1.3) |
| **Setting** |  |
| ICU setting | 14 (18.4) |
| Other hospital setting | 52 (68.4) |
| Primary care | 6 (7.9) |
| Other healthcare service | 2 (2.6) |
| *Missing* | 2 (2.6) |

### Accessibility and implementation of CMGs

Most of the respondents (67%) reported having used a Covid-19 CMG to guide clinical decision-making within the last two weeks. A majority of the respondents, 87% (44/51) used a local or national CMG, and 27% (14/51) also the WHO CMG to guide clinical decision making. About a third (38%, 27/73) reported a need for additional or updated CMGs, particularly for critically ill people in hospital (96%), people treated at home (73%), children and for pregnant women. Just over half (57%, 40/70) of respondents agreed or strongly agreed that they had received sufficient training in caring for Covid-19 patients and 77% (43/65) that they felt confident in implementing the recommendations in the available Covid-19 CMGs, but only 58% (41/71) to having access to the treatment and equipment needed to implement the recommendations. This proportion was higher amongst respondents based in LMICs, reporting limited access to oxygen, ventilators, observational, specimen collection and personal protective equipment (Fig 3).

**Figure 3.**
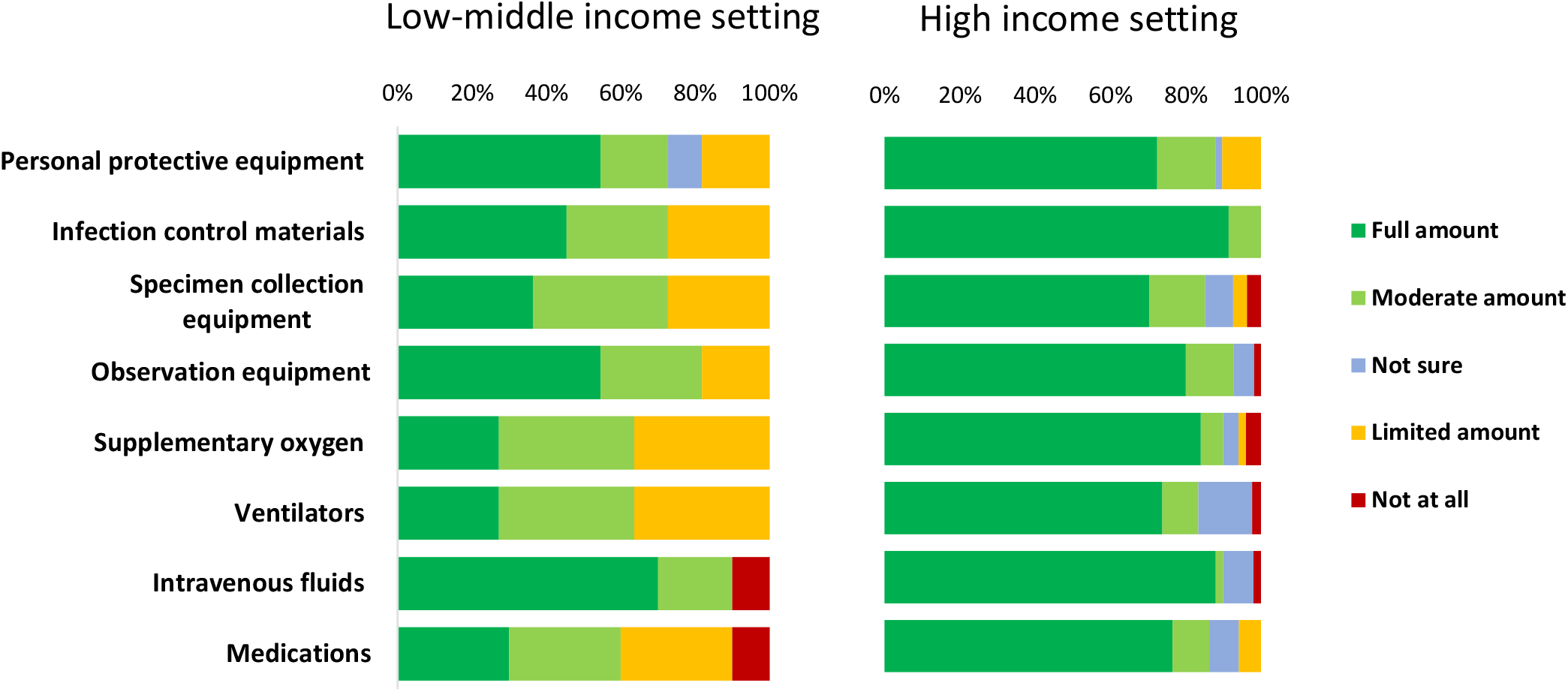
Access to treatment and equipment for Covid-19 clinical care. The figure presents the proportions of respondents reporting different level of access to Covid-19 treatment and equipment at their site.

### Inclusivity of CMGs

The responses show good access to CMGs providing recommendations for adults, but limited access to evidence-based recommendations for other risk groups, with only 20% (15/76) of respondents reporting having access to CMGs that covered guidance on treatment decisions for children, 25% for pregnant women, 32% for people who are immunosuppressed and 50% for older adults (>65) years presenting with suspected or confirmed COVID-19. None of the respondents from LMIC reported having access to CMGs for care of children or pregnant women with Covid-19 (Fig.4).

**Figure 4.**
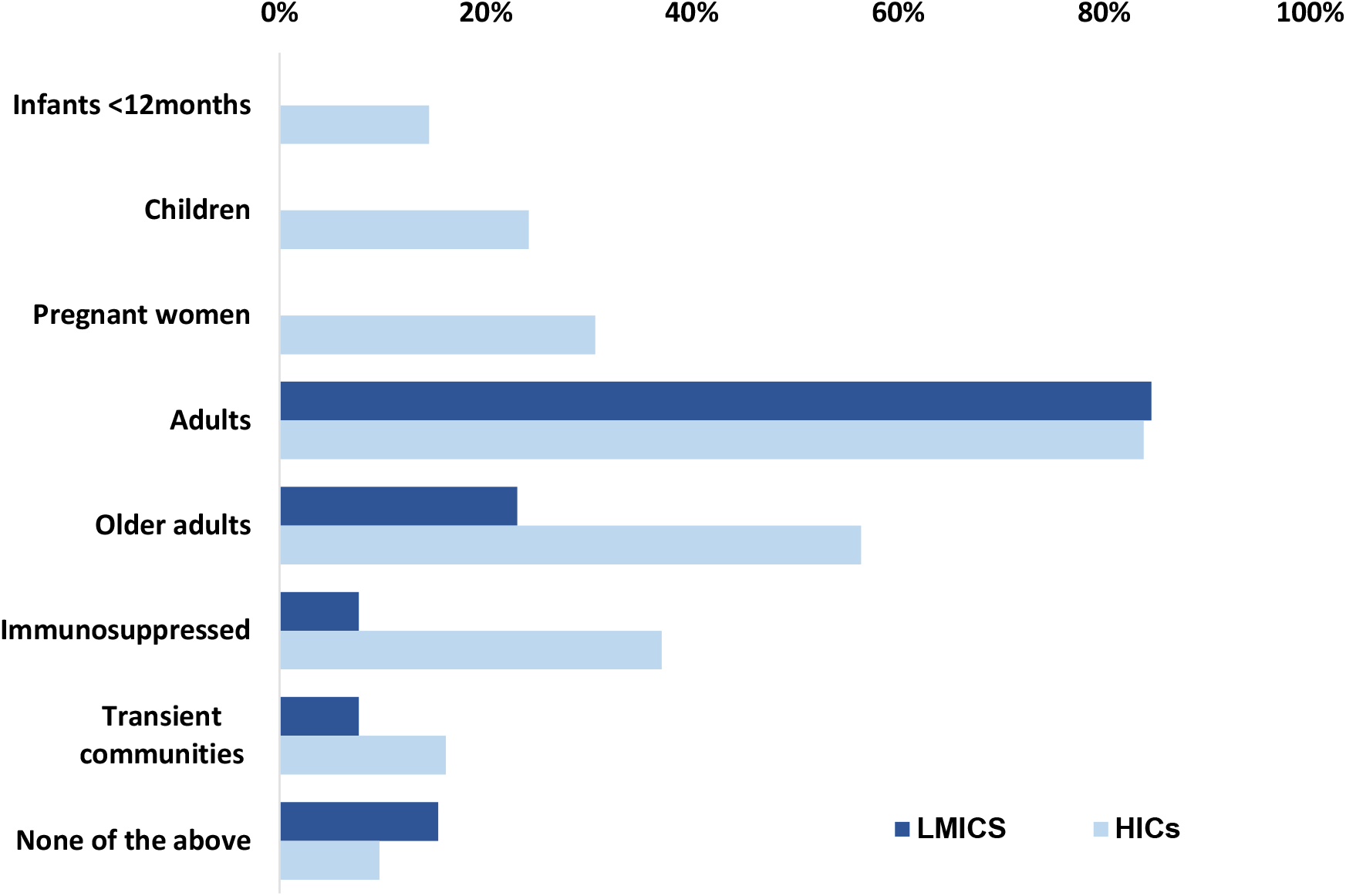
Access to recommendations for different at-risk populations. The proportion of respondents reporting that the available Covid-19 CMGs includes evidence-based treatment recommendations for different at-risk populations by different resourced settings. **Abbreviations:** CMGs: Clinical management guidelines; LMICs: low-middle income countries, HICs: high income countries

### Recommendations on improvements of CMG development

Themes that were identified from the respondents were a need to harmonize international CMG development, to use resources effectively, minimize guidelines variations, and during uncertainty and limited evidence, formulate recommendations through international consensus. Guideline transparency was recommended, highlighting where the evidence base is limited or uncertain. Many respondents called for CMGs to include recommendations for different at-risk groups, such as children, and to be applicable to lower resourced settings. There were also calls for supportive care and treatment recommendations to be more comprehensive, and cover follow up care. Importantly, implementation of CMGs needs to be supported with sufficient training, access to recommended treatment as well as personal protective equipment for staff. Moreover, to facilitate implementation, CMGs should be brief and supported with clear flowcharts. Some respondents reported that the available CMGs were out of date, a living guideline approach, for readily updating CMGs as new evidence emerges was recommended.

## Discussion

These findings highlight crucial areas for improvement when developing infectious disease CMGs for emerging pathogens during public health emergencies. Our data shows that despite good access to CMGs for COVID-19, there were gaps in scope, applicability and inclusivity, including limited access to CMGs containing recommendations for children, pregnant women and older people. For COVID-19 and other infectious diseases, these groups may present different risks regarding epidemiology, severity and complications, and may present with different symptomatology, which must be considered for timely identification and diagnosis and optimal management and treatment strategies. Similar gaps in scope and inclusivity have been identified in systematic reviews of CMGs for other viral infectious diseases. These vulnerable risk groups may be disproportionately affected in health crises (20), and require care appropriate to their needs throughout the response to safeguard against potentially avoidable deaths (11). Further, this survey identified that limited access to evidence based recommended medical treatments and equipment was a critical barrier to the implementation of available COVID-19 CMGs, especially affecting LMIC settings. This highlights the discordance between guideline development, often undertaken by stakeholders in HICs (2), and clinical practice in varied contexts subject to suboptimal resource availability. Further, the lack of attention to considering the realities of guideline implementation, especially in low resource contexts.

Our data also highlights that during an emergency frontline staff may have to task-shift, support other specialties and step up to take on new responsibilities. Clinicians rely on CMGS to turn the best available evidence into clinical practice to improve patient outcomes. To do so requires ready access to high-quality guidelines applicable to relevant patient groups and contexts. In emergencies, where time and evidence are limited, strict guideline development standards may seem inflexible and unnecessary. Nevertheless, clinicians must trust that available CMGs are evidence-based, as well as relevant to implementation in their local context. Our data demonstrate that directly transferring CMGs between contexts, specifically those developed in HICs to LMICs, may not always be appropriate or effective. The COVID-19 pandemic has generated new evidence at unprecedented speed, highlighting that CMG developers need to consider tools for rapidly updating and re-disseminating CMGs as new evidence emerges.

Whilst there is a definite role for leadership bodies that can widely disseminate guidelines, well-resourced organisations such as WHO need to engage diverse stakeholders from different resourced settings to ensure that CMGs address the needs of different risk populations. A previous study showed that fewer guidelines and less locally-relevant trial evidence are produced in LMICs (3). Other studies have shown that local guidelines developed in collaboration with local stakeholders are more likely to be effectively implemented, as these better account for available resources, specialist skills, and cultural influences; thus promoting ownership and focusing on specific contextual needs (3,21). As Atkins et al. (22) put it: “Evidence does not form recommendations on its own.” Guideline development requires multiple inputs beyond scientific evidence, including expert opinion and previous experiences, especially for new emerging diseases like COVID-19 where evidence is limited. Social science literature further emphasises the interaction of these aspects, and the importance of understanding the social processes of how guideline development committees with different backgrounds and priorities interact and communicate, influencing how evidence is considered (23).

*De novo* guideline development is resource-intensive and all setting may not have the resources to develop CMGs during a novel epidemic or pandemic, yet more adaptable approaches require existing CMGs to be high-quality and up-to-date (24). Following Dagens *et al*. (2), a ‘living guidelines’ approach as recently adopted by WHO may better adapt to rapidly evolving emerging outbreaks (25,26). This should include a flexible tiered approach, with recommendations tailored to different resourced health systems, enabling sites to switch between tiers as resources becomes available or depleted. Local prioritisation processes in the form of rapid research needs appraisals could also identify key contextual areas to update (27,28).

Whilst the survey was disseminated widely, the response rate was limited by the ongoing pandemic response. Despite these limitations, the responses represent a wide range of settings globally, and identifies critical gaps and a unique cross-sectional insight into the access, inclusivity, and implementation of CMGs early on during a pandemic response.

This study highlights gaps in inclusivity of and access to COVID-19 CMGs for different resourced settings and populations, and challenges in implementation during an emergency. The findings highlight a need for a new evidence-based framework for infectious diseases CMG development. This framework should include stakeholders from different resourced settings, to ensure locally relevant challenges and priorities are addressed. CMGs needs to be flexible, and adaptable to new emerging evidence and situations, considering implementation support for different settings. A living review framework addressing implementation facilitators were recommended. A failure to invest in an updated framework for infectious disease CMG development risks widening health inequalities.

## Data Availability

The full dataset generated and analysed during the current study is available from the corresponding author on reasonable request.

## Acknowledgments

ISARIC members, PREPARE colleagues, and clinicians worldwide that responded to the survey. Dr. Terrence Epie and Dr. Mais Tattan for piloting and disseminating the survey through local networks in sub-Saharan Africa and the Middle East. Sarah Moore, Romans Matulevics and Melina Michelen in the ISARIC Global Support Centre for invaluable administrative support.

## Authors’ contributions

Conceptualisation and design of the project (LS, CP, DM, KN, NG), survey design and piloting (LS, CP, DM, KN, NG), survey programming (KK), survey dissemination (KK, LS, CP, DM, KN), data analysis and interpretation (LS, CP, DM, KN, KK, AD), manuscript writing (CP, LS, DM, KN, AD, MM). All authors reviewed and approved the final manuscript.

## Ethics approval and consent to participate

All participants were given online information about the study, and the opportunity to ask questions via email of the authors, before completing the survey. Informed consent was given by all participants through their completion of the survey online.

## Consent for publication

Not applicable

---

**Supplemental file 1.** Covid-19 CMG Survey.

| Variable | Format options |
| --- | --- |
| Age | Range: 18 – 24, 25-34, 35-44, 45-54, 55-64, 65 - 70, >70 , Prefer not to say |
| Gender | Female, Male, Non-binary, Prefer not to say |
| Country of work (if more than one, select the one currently/mainly working in) |  |
| Job role | Healthcare assistant, Nurse, Paramedic, Senior medical doctor, junior medical doctor, Other (specify) |
| Type of specialty (specify): |  |
| Type of healthcare service* (tick all that apply): | Primary care, Hospital, Accident and Emergency Unit, Intensive care unit, Rehabilitation, Palliative care unit, Care Home/Nursing Home, other: Specify:) |
| Are you currently or have you been working in a different clinical role in order to support the COVID-19 response? | Yes/ No<br>If yes, please specify: |
| “During the COVID-19 pandemic I have been taking on a higher level of responsibility than would normally be expected of me” | Yes/No/NOT SURE |
| Over the past 2 weeks, have you personally provided treatment or care for patient(s) with suspected or confirmed COVID-19 infection in a clinical setting? | Yes /No / N/A |

## Please identify the extent to which you agree or not with the following statements

Response option Q2-4: 7 point Likert scale: strongly disagree – disagree – somewhat disagree – neither agree nor disagree – somewhat agree – agree – strongly agree or N/A

## Implementation indicators

1a. Over the past 2 weeks, I have used clinical management guidelines to guide my clinical decision-making on care and treatment to patients with suspected or confirmed COVID-19. Yes / No /N/A IF YES: Which organization developed these guidelines? (Tick all that apply)
  - Local hospital
  - Local public health
  - National public health agency
  - A Centre for Disease control (e.g. US CDC, ECDC, Africa CDC, China CDC)
  - WHO
  - Other organization: (please specify):
1b. In your setting, Is there a need for additional or updated clinical management guidelines to guide care and treatment decisions for people with COVID-19 patients? Yes No Not Sure. If yes, please list type of clinical guidelines needed (tick all):
  - For care of people with COVID-19 in hospital
  - For care of people with mild COVID-19 at home
  - For care of more severely ill people with COVID-19 at home (including palliative care)
  - For care of people with COVID-19 in primary care
  - For care of people with COVID-19 in care homes
  - For care of people with COVID-19 in ICU
  - For care of children with COVID-19
  - Other (specify):
2. I feel I have received sufficient training in the clinical management of patients presenting with suspected or confirmed COVID-19.
3. Over the past 2 weeks, I have felt confident in following the information provided in written guidelines to inform my clinical decision-making about providing care and treatment to patients with suspected or confirmed COVID-19.
4. I can easily access the equipment and materials needed to follow recommended guidelines on treatment of patients with suspected or confirmed COVID-19.
5. Over the past week, to what extent were the following materials for COVID-19 clinical management available? [Response options: not at all – limited amount – moderate amount – full amount – Not Sure - Not applicable]
  - Personal protective equipment e.g. surgical masks, gloves, eye shields, goggles etc.
  - Infection control materials e.g. alcohol hand wash, soap, running water, disinfectant
  - Specimen/sample collection equipment – e.g. swabs
  - Observation equipment e.g. temperature probe, Blood pressure (BP) cuff, Oxygen Saturation probe
  - Supplementary oxygen: e.g. Nasal oxygen, Venturi mask oxygen, CPAP, HFNO, IN ITU
  - Ventilators (if part of your practice), In-line suction catheters, Capability to undertake prone positioning, Neuromuscular blockade drugs, Vasopressors
  - Intravenous fluids including cannulation equipment
  - Medications, including antivirals if recommended and empirical antibiotics where needed
  - Other

## Views on guideline availability, credibility, and inclusivity

Response option Q7 – 8 and10: 7 point Likert scale: strongly disagree – disagree – somewhat disagree – neither agree nor disagree – somewhat agree – agree – strongly agree or N/A

3. The available written or online clinical management guidelines to guide treatment decisions for patients with suspected or confirmed COVID-19 are easily available.
4. The available written or online clinical management guidelines to guide treatment decisions for patients with COVID-19 are evidence-based.
5. The available written clinical management guidelines to guide treatment decisions for patients with suspected or confirmed COVID-19 sufficiently address the needs of (tick all that apply) – drop down list of options:
  a. Infants <12 months of age
  b. Children
  c. Pregnant women,
  d. Older adults (over 65yrs)
  e. Frail older adults
  f. Those who are immunosuppressed,
  g. Those with comorbidities,
  h. Transient communities (e.g. traveller, migrants).
6. Please mention any other comments on how the clinical management guidelines for COVID-19 could be improved:

